# Enhancing Sweat Rate for In-Hospital and Home-Based Decongestive Therapy

**DOI:** 10.1101/2024.05.05.24306883

**Authors:** Doron Aronson, Yaacov Nitzan, Sirouch Petcherski, Aviv Shaul, William T. Abraham, Daniel Burkhoff, Tuvia Ben Gal

**Affiliations:** Departments of Cardiology, Rambam Medical Center, and B. Rappaport Faculty of Medicine, Technion Medical School, Haifa, Israel; AquaPass Ltd.; Cardiovascular Research Foundation, New York, New York; Division of Cardiovascular Medicine, The Ohio State University, Columbus, Ohio; Heart Failure Unit, Cardiology Department, Rabin Medical Center, Faculty of Medicine, Tel Aviv University, Tel Aviv, Israel

**Keywords:** Congestion, Diuretic response, Edema, Heart failure, Sweat, Volume overload

## Abstract

**Background:** Many patients admitted for acutely decompensated heart failure (ADHF) are discharged with persistent congestion. Furthermore, the early post-discharge period carries a particularly high risk of readmission and is known as the ‘vulnerable phase’.

**Methods:** We used a device designed to enhance fluid and salt expulsion via the eccrine sweat glands for decongestion of patients admitted for ADHF. Patients were treated for 1-6 days in the hospital. Following discharge, home therapy continued for 30-60 days at a rate of 1-4 treatments/week. The primary efficacy endpoint for the in-hospital phase was fluid loss of ≥500 mL per ≥4h treatment. Secondary performance endpoints including fluid loss, changes in congestion score, and changes in NT-pro-BNP levels were evaluated for each phase separately.

**Results:** We studied 15 patients, 12 completing both the hospital and home phases. During the in-hospital and home phases, median weight change due to device therapy was 2.4 Kg [IQR 2.20–3.77] and 3.1Kg [IQR 7.4–0.6 Kg] respectively, and the primary endpoint was met in 86% of patients. During the home treatment, median weight loss was 3.1 Kg [IQR 7.4–0.6 Kg]. Congestion score declined from 6 [IQR 6 to 7] to 4 [IQR 3–6] during the hospital phase (P=0.0007) and to 1 [IQR 1–1.5] at the end of home therapy (P=0.002). Median NT-proBNP levels decreased from 7732 [IQR 4694– 9746] to 4984 pg/mL [IQR 3559–8950](*P*=0.01) during the hospital phase and to 3596 ng/mL [IQR 1640–5742](*P*=0.02) at the end of home therapy. Renal function remained stable.

**Conclusion:** The AquaPass device is useful in enhancing decongestion in hospitalized ADHF patients. Following hospital discharge, device therapy was associated with additional improvement in decongestion without worsening of renal function.

## Introduction

Acute decompensated heart failure (ADHF) is a major public health problem, culminating in more than 1 million annual hospitalizations in the United States alone ^1^. Most patients who are hospitalized with worsening heart failure do not have a new, acute disorder. Rather, they suffer from chronic heart failure and present in a decompensated state as a consequence of gradual but progressive fluid accumulation and increases of cardiac filling pressures (both central venous and pulmonary capillary wedge pressure) over the preceding weeks ^2, 3^

Despite modern therapy, a significant proportion of patients admitted with a primary diagnosis of ADHF experience minimal weight loss or may even experience weight gain during hospitalization.^4–6^ Furthermore, the early (first 2–3 months) post-discharge period immediately following hospitalization, carries a particularly high risk of poor clinical outcome and is known as the ‘vulnerable phase’.^7^ The pathophysiology underlying these early adverse events is likely associated with persistent congestion at the time of discharge and subsequent and/or a gradual re-accumulation of fluids after discharge.^8, 9^ However, maintaining optimal volume status in the early post-discharge period remains a challenge. Consequently, decongestion in the post-hospital discharge outpatient setting represents an unmet need to improve quality of life and reduce rehospitalizations.^10–12^

The mainstay of therapy targeting hypervolemia and congestion is the use of loop diuretics. We have recently described a novel strategy for fluid removal by increasing sweat rate via activation of the eccrine glands, thus removing fluids and sodium directly from the interstitial compartment as a potential treatment of congestion in heart failure.^13^ The aim of the present study was to test the feasibility and efficacy of this approach as an additional decongestive therapy in hospitalized ADHF. We also studied the effects of this therapy on indices of congestion in the early post hospital discharge period.

## Methods

The study was conducted under the approval of the institutional review boards at the Rambam Medical Center, Haifa, Israel and the Rabin Medical Center, Petah Tikva, Israel. Written informed consent was obtained from all study participants prior to study procedures. The protocol was registered at ClinicalTrials.gov (Unique identifier: NCT05843201).

### Inclusion and exclusion criteria

This study recruited patients hospitalized for worsened heart failure and evidence of persistent fluid overload (as evidenced by a modified EVEREST composite congestion score ≥3)^6^ despite receiving ≥80 mg of furosemide IV per day during the index hospitalization, age ≥18 years, estimated glomerular filtration rate >15 ml/min/1.73 m^2^ (calculated based on the Chronic Kidney Disease Epidemiology Collaboration (CKD-EPI) equation)^14^, baseline systolic blood pressure ≥100 mmHg, and elevated NT-proBNP (NT-proBNP >1,600 pg/ml for BMI <30 kg/m**^2^**; NT-proBNP >800 pg/ml patients with BMI >30 kg/m**^2^**; NT-proBNP >2,400 pg/ml for patients with persistent or permanent atrial fibrillation).

It was also required that patients complete a 2-hour run-in acclimatization session with the AquaPass system (described below) to demonstrate adequate treatment-induced sweat response (defined as ≥100 mL/hr) and confirm the patients’ willingness to participate in full, 4-hour treatment sessions.

Exclusion criteria included: 1) lower extremity skin condition (open wounds, ulcers); 2) severe peripheral arterial disease; 3) eGFR<15 ml/min/1.73 m**^2^** or requiring dialysis; 4) receiving pressors, inotropes, mechanical circulatory support or mechanical ventilation; 5) experiencing frequent or sustained cardiac arrythmias (other than chronic atrial fibrillation); 6) active infection; 7) history of heart transplant, actively listed for heart transplant or planned left ventricular assist device; 8) malignancy or other noncardiac condition limiting life expectancy to <12 months.

### Study protocol

This study was divided into in-hospital and out-of-hospital phases. For the in-hospital phase, patients were initially treated by the hospital physician with diuretic therapy in a manner consistent with European Society of Cardiology^15^ and AHA/ACC/HFSA guidelines.^16^ Patients with persistent congestion and meeting all study entry criteria were offered participation in this study. After providing informed consent, patients underwent daily AquaPass treatment, with each treatment lasting up to 4 hours (up to 5 treatments). Diuretic therapies remained constant during this phase unless dictated otherwise by patient conditions. Daily measurements included body weight, creatinine, BUN, serum electrolytes, and NT-proBNP. Decisions regarding hospital discharge where at the discretion of the treating physician.

After completion of the in-hospital phase, patients were provided a second informed consent to enter the out-of-hospital therapy phase of the study. AquaPass treatments were performed at home, initially at 3 times per week, with each session lasting ∼4 hours. AquaPass treatment frequency was modified at the discretion of the study investigators according to patient condition and ranged from 1 to 4 sessions per week for between 4 and 8 weeks including the in- and out-of-hospital study phases. Similarly, diuretic therapy was also varied by the study investigator based on the degree of residual congestion.

### The AquaPass System

The AquaPass system (AquaPass Ltd., Shefayim, Israel), designed to enhance fluid and salt removal via the eccrine sweat glands, has been described previously.^13^ Briefly, the system is comprised of two main functional units: (1) a wearable garment, and (2) a control station with a heating unit and fan. The garment creates a homogeneous warm temperature environment around the lower part of the body leading to increased skin temperature that activates the eccrine glands and initiates perspiration. The sweat evaporates instantaneously, thus avoiding the awareness of perspiration by the patient and enabling long durations of treatments, if required.

The heating sub-unit controls the temperature inside the garment. The skin temperature is uniformly increased from 32-33□ to the range of 36–40□. Previous studies have shown that the relationship between skin temperature and sweat production is linear^17–19^ and that neither discomfort nor thermal injury occur.^20^ The median hourly weight loss induced by the device is ∼200 g.^13^

A user interface enables the operator to set the air flow rate and maintain skin temperature within a user-specified range of values. During the treatment, measurements made by the sensors in the wearable garment and the calculated sweat rate are displayed on the console.

### Study endpoints

The primary performance endpoint for the in-hospital phase was the amount of fluid loss from sweat per treatment procedure; the target rate of sweat production was >500 ml/session. Other exploratory endpoints for the in-hospital phase included: 1) changes of clinical congestion score; 2) the need to increase diuretic dose by ≥50% compared with the initial dose; 3) change in NT-pro-BNP; 4) cumulative loop diuretic dose during index hospitalization; and 5) changes in renal function assessed by serum creatinine and BUN.

Exploratory performance endpoints assessed during the out-of-hospital, home therapy study phase consisted of within-subject differences from discharge to end of treatment values in: 1) fluid loss due to AquaPass treatments; 2) change of diuretic therapy (quantified by daily equivalent dose of furosemide); 3) change in congestion score; 4) change in body weight; 5) change in quality of life as assessed by the Kansas City Cardiac Questionnaire-12 (KCCQ-12); 5) change in NT-pro-BNP; 6) change in serum creatinine; and 7) the incidence of hospitalization or emergency department visits for heart failure.

### Statistical analysis

Categorical variables are presented as frequencies. Continuous variables are presented as mean ± SD or median with interquartile range (IQR). Continuous variables were tested for normal distribution using histograms and quantile-quantile plots.

Differences in continuous variables between different time points were analyzed using paired-sample Student *t*-tests or Wilcoxon matched-pairs singed rank tests (for within-group differences), and the Mann-Whitney *U* test (for between-group differences).

For repeated measurements, we applied linear mixed models in order to account for intra-subject correlation,^21^ with an unstructured covariance matrix and small sample adjustment made using the Kenward□Roger method.^22^ Differences were considered statistically significant at the 2-sided *P*< 0.05 level. Statistical analyses were performed using the Stata version 18.0 (College station, TX).

## Results

Between February 2022 and July 2023, 15 patients with ADHF (mean age, 70±13 years; 14 male) were recruited into the study. Of these, 12 patients signed the additional informed consent for home therapy. The clinical characteristics of the treated patients are summarized in Table 1.

**TABLE 1:** Characteristics of the heart failure patients (N=15)

| Characteristic |  |
| --- | --- |
| Age | 72 ± 8 |
| Male | 14 (93%) |
| Hypertension | 12 (8%) |
| Diabetes | 8 (53%) |
| NYHA functional class |  |
| III | 8 (53%) |
| IV | 7 (47%) |
| Ischemic heart disease | 10 (66%) |
| Atrial fibrillation | 11 (73%) |
| ICD/CRT | 5 (33%) |
| Baseline creatinine (mg/dl) | 2.10 [1.09, 2.46] |
| Estimated GFR (ml·min<sup>-1</sup>/1.73 m<sup>2</sup>) | 35 [26, 68] |
| Blood urea nitrogen (mg/dL) | 43 [33, 74] |
| Hemoglobin (g/dL) | 11.1 ± 1.6 |
| Sodium (mmol/L) | 140 ± 3 |
| Potassium (mmol/L) | 4.0 ± 0.6 |
| Urine sodium (meq/L) | 62 [44, 82] |
| NT-proBNP (pg/mL) | 7732 [5016, 11704] |
| LVEF (%) | 37 ± 20 |
| Pulmonary artery systolic pressure (mm Hg) | 51 [41, 58] |
| Beta blockers | 11 (85%) |
| ACE Inhibitors/ARB/Scubitril-vasartan | 10 (69%) |
| MRA | 13 (87) |

**TABLE 2:** Renal function and electrolytes.

|  | **In-Hospital (n=48)** | **Home Therapy (n=48)** | ***P* Value** |
| --- | --- | --- | --- |
| **Creatinine (mg/dL)** | 1.66 [1.25–2.00] | 1.63 [1.20–2.06] | 0.82 |
| **BUN (mg/mL)** | 26 [18–48] | 25 [16–46] | 0.14 |
| **Serum Na (mmol/L)** | 139 ± 2 | 140 ± 2 | 0.002 |
| **Serum K (mmol/L)** | 4.7 ± 0.4 | 4.6 ± 0.4 | 0.56 |
| **Urine Na (mmol/L)** | 50 [31–66] | 45 [20–72] | 0.49 |

Comparisons are for each treatment session

### In hospital study phase

The mean number of treatments during hospitalization was 4.2 (range 1 to 6) per patient. Overall, a total of 51 procedures were completed. The patients were concomitantly treated with diuretics with a median daily furosemide dose of 160 mg/day [IQR 120 to 240 mg/day]).

During the 51 treatment sessions, fluid loss from the device had a median value of 166 gr/hr (IQR, 134–225 gr/hr). The median total weight change per treatment session (after accounting for urine output, fluid intake and food consumed during the procedures) was a net reduction of 678 g (IQR, 578–885 g). Sweat exceeded 500 gr over the 4-hour treatment in 44 of the 51 (86%) sessions. Overall, on days of AquaPass treatments, the mean daily weight loss (i.e., over 24 hours) was 1.2 Kg (95% CI, 0.9 to 1.5 Kg). Cumulative sweat loss over all in-hospital Aquapass treatments had a median value of 2.4 Kg per patient [IQR 2.20 to 3.67 Kg]. Body weight decreased by 2.5 ± 3.2 Kg (range -11.0 to 0.2 Kg) (*P*<0.5) during the in-hospital study period.

Congestion score had a median value of 6 [IQR 6 to 7] at baseline and decreased to 4 [IQR 3 to 4] (*P*=0.0007, Figure 1) at hospital discharge. NT-proBNP decreased significantly as summarized in Figure 2 (p=0.01).

**Figure 1:**
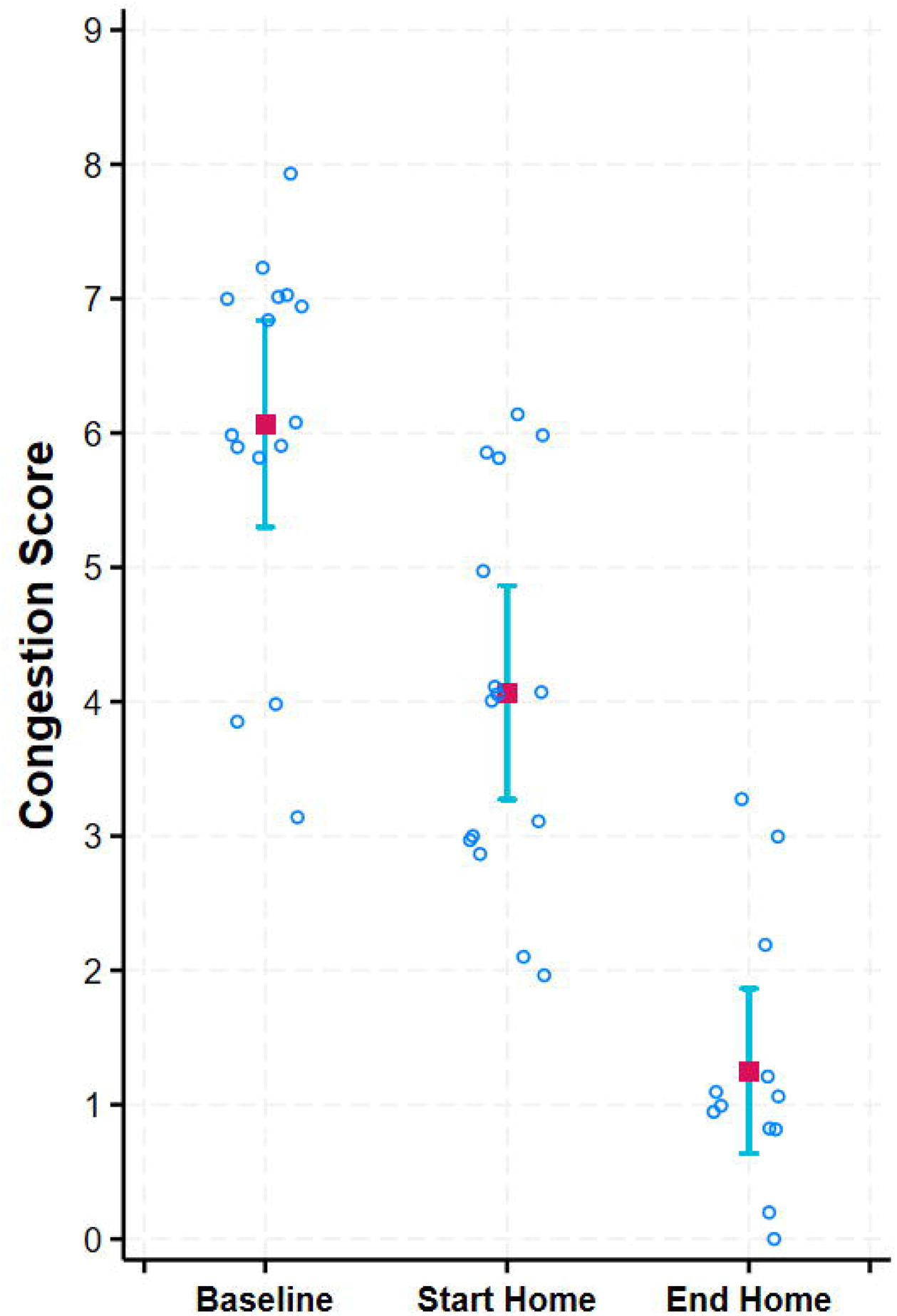
Changes in congestion score with (A) hospital therapy (n=15) and (B) Home therapy (n = 12).

**Figure 2:**
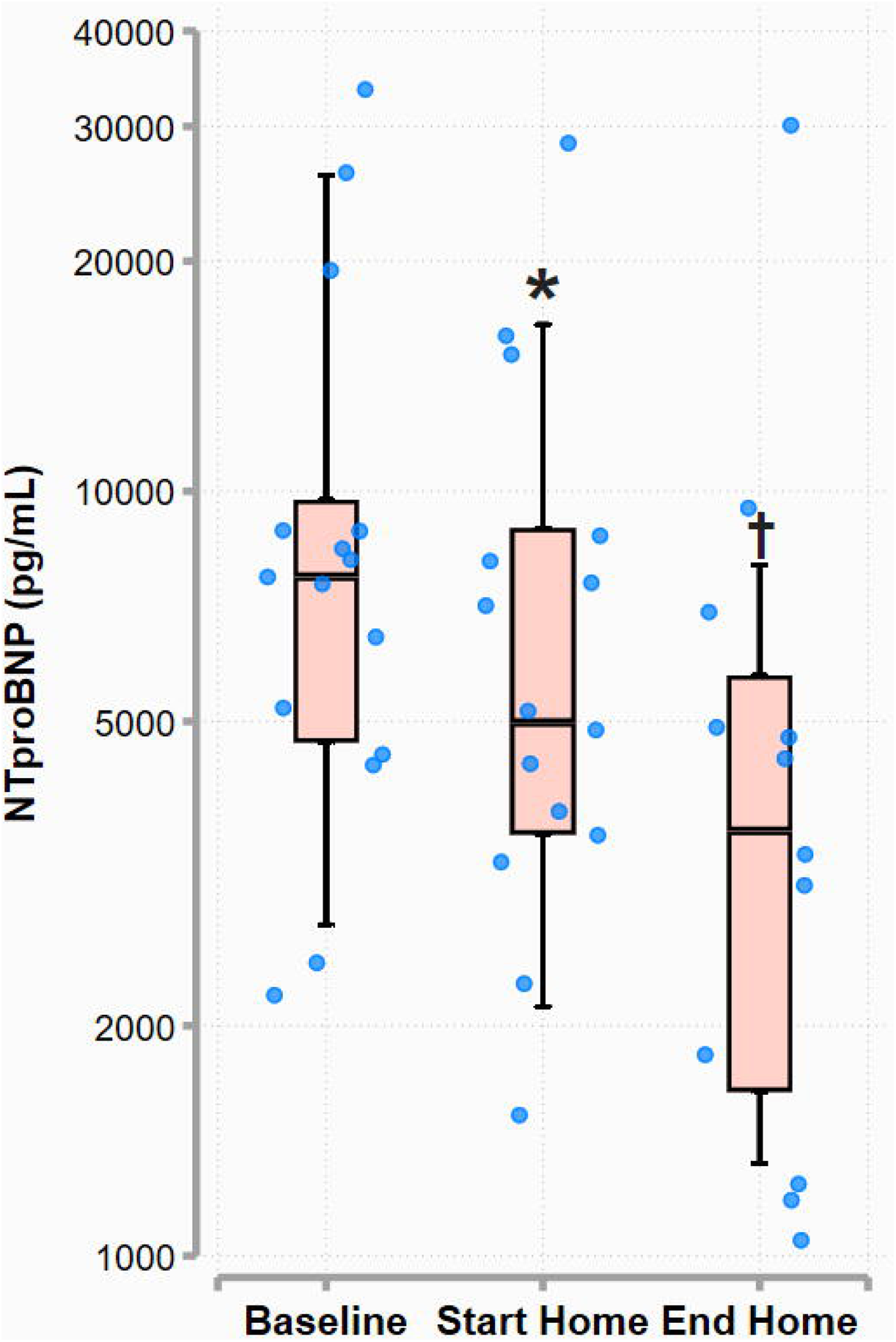
Changes in NT-proBNP levels during the study.

### Outpatient phase

Twelve patients continued with home therapy after hospital discharge. These patients received 5 to 18 home treatments (median 14 treatments) over a median of 34 days (IQR 28 to 39 days). The median total weight reduction per each home treatment session was 806 gr (IQR, 627–1003 gr; hourly median weight loss from sweat 195 gr/h; IQR, 152– 225). For a typical 3 treatments per week, this culminated in a median fluid removal of ∼2.4 L/week. Sweat rates were maintained from one session to the other over the course of the study (Supplemental Figure 1; *P* for linear trend across treatment sessions *P*=0.06). During the 34 median days of the out-patient phase of the study, 10 of 12 patients lost weight, with a median weight loss of 3.1 Kg [IQR 7.4 to 0.6 Kg].

Concomitant with this weight loss, the congestion score declined from 4 [IQR 3 to 6] at the end of hospital therapy to 1 [IQR 1 to 1.5] at the end of home therapy (*p*=0.002; Figure 1). NT-proBNP levels decreased during home therapy from 4984 pg/mL [IQR 3559 to 8950 pg/mL] at discharge to 3596 ng/mL [IQR 1640 to 5742 pg/mL] at the end of home therapy (Figure 2, *P* = 0.02). The median weekly furosemide dose decreased from 1190 mg/week [IQR 960–1540] at hospital discharge to 980 mg/week [700–1120] (*P*=0.01) at the last study week (Supplemental Figure 2). A reduction in oral furosemide dose of ≥25% occurred in 6 patients. Sweat rate was independent of renal function. Median hourly sweat rates averaged 156 [IQR 107–186] gr, 143 [IQR 86–170] gr and 176 [IQR 121–214] gr in patients with eGFRs of 15-29, 30-59, and ≥ 60 ml·min^-1^/1.73 m^2^, respectively (Supplemental Figure 3, *P*=0.11).

The median improvement in heart-failure symptoms from baseline to the beginning of home therapy was 14.3% (IQR, 1.3 to 38.9%; *P*=0.01 compared with baseline) as measured on the KCCQ-12. During home therapy, the KCCQ-12 increased further by a median of 24.0% (IQR, 15.1 to 27.5%, *P*=0.004) (Figure 3).

**Figure 3:**
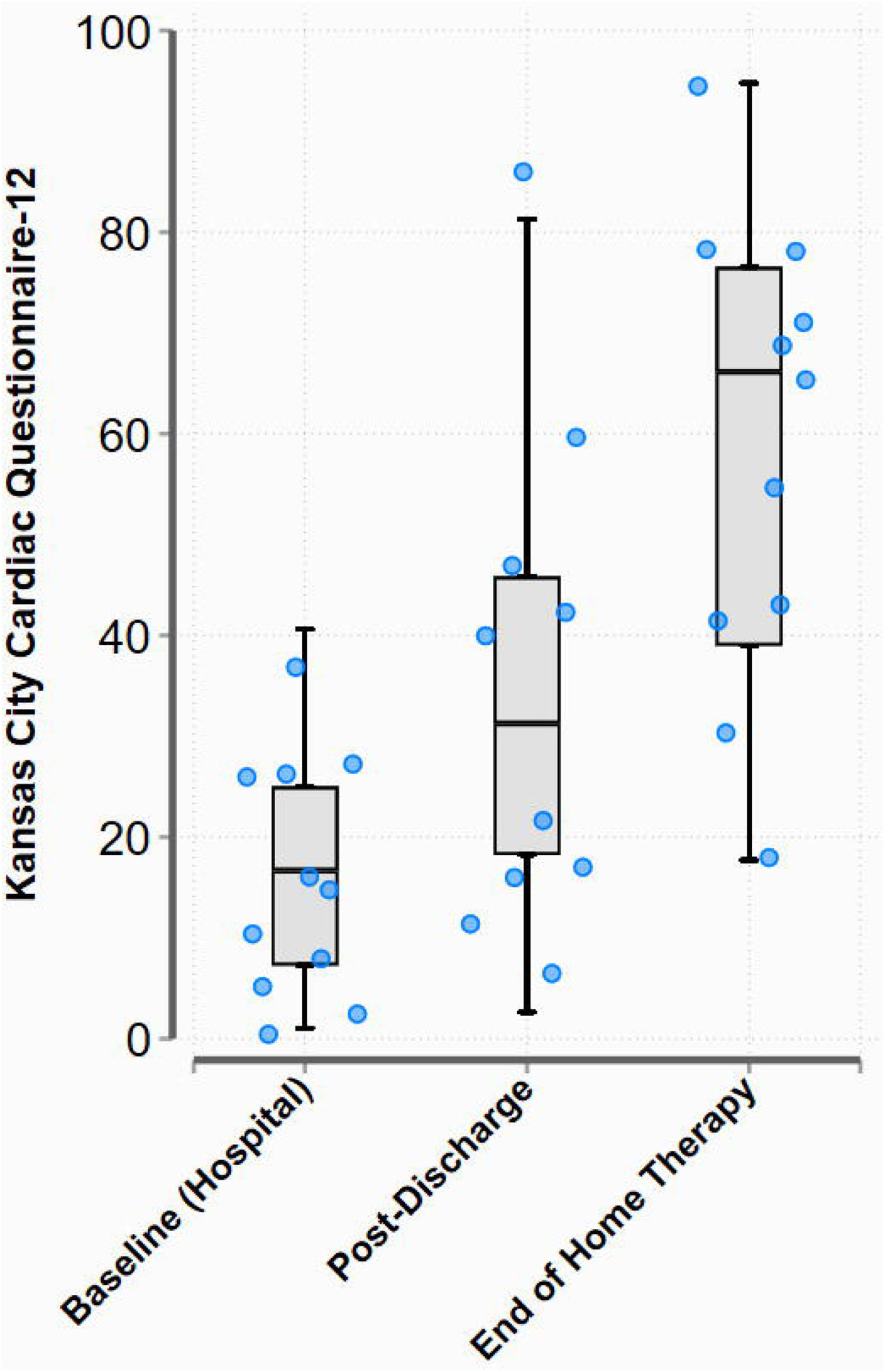
Overall summary score on the Kansas City Cardiomyopathy Questionnaire (KCCQ-12) at baseline, start of home therapy and end of home therapy. **\*** *P*<0.01; **^†^** *P*<0.05

Figure 4 summarizes changes in serum creatinine from baseline to end of the study in all participants. There was no significant change in renal function over the course of the study (baseline creatinine 1.73 ± 0.18 mg/dL vs. ending creatinine 1.72 ± 0.19 mg/dL, *P*=0.84). None of the patients was readmitted for HF.

**Figure 4:**
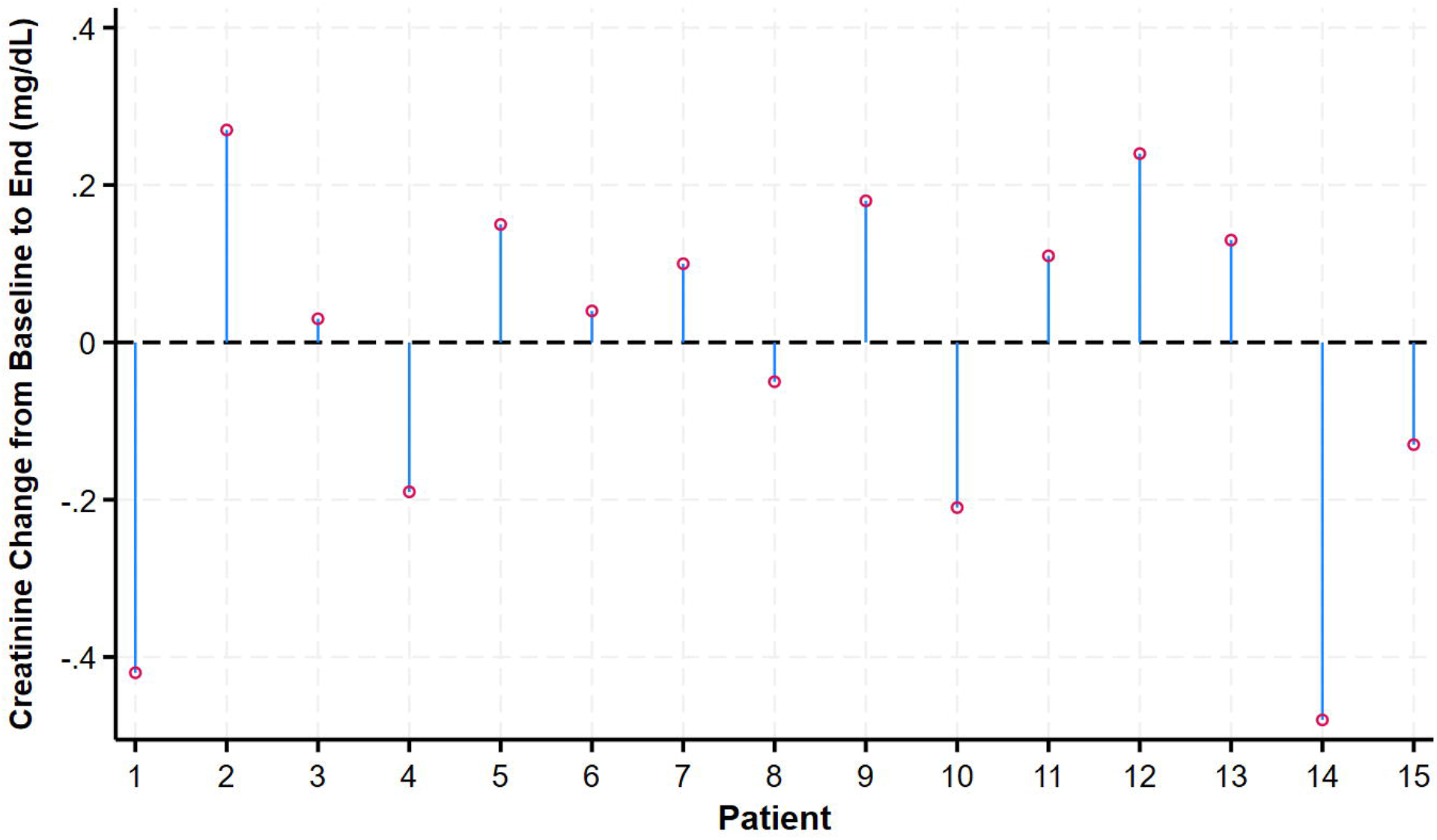
Changes in serum creatinine during the study in all patients. Red circles represent the change in serum creatinine from baseline to the end of the study.

Figure 5 displays the blood pressure changes during the procedures. The largest decline in systolic and diastolic blood pressure occurred at the 2^nd^ or 3^rd^ hour timepoint [-4.3 mm Hg (95% CI, -6.0 to -2.6) and -3.1 mm Hg (95% CI, -4.5 to -1.7), respectively]. Heart rate increased slightly, peaking at the 4^th^ hour of the procedure (1.9 BPM; 95% CI, 0.7 to 3.1).

**Figure 5:**
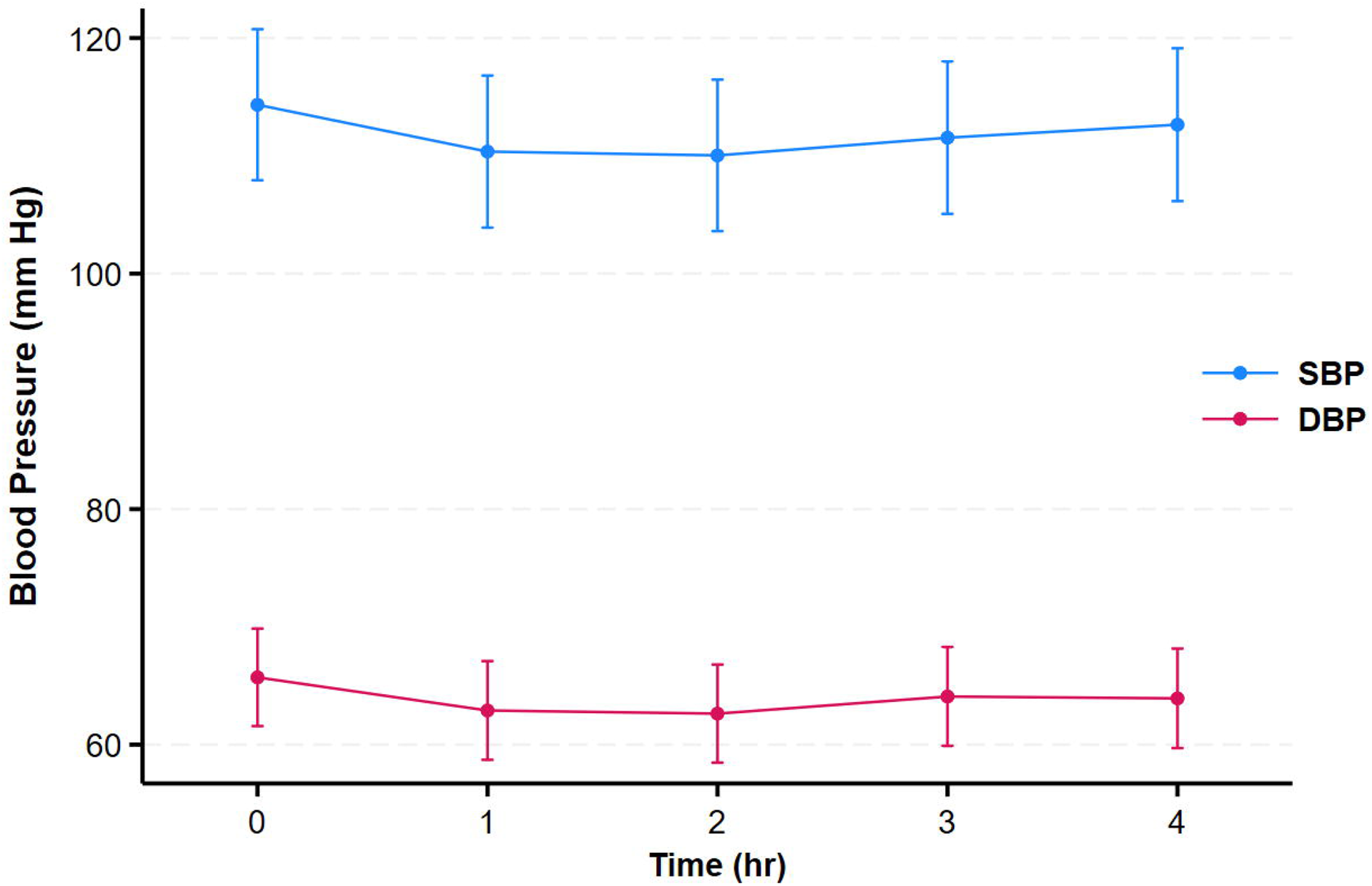
Mean (95% CI) changes in systolic and diastolic blood pressure during the procedures.

## Discussion

The present study tested the safety and feasibility of the AquaPass system as a method to enhance decongestion in hospitalized, fluid overloaded patients, safely continue decongestion as an outpatient, and prevent fluid re-accumulation in the vulnerable post-discharge period with home therapy. During in-hospital therapy, the device provided clinically meaningful rates of fluid removal. During the home therapy phase, the therapy resulted in further fluid removal and reductions in congestion score and NT-proBNP levels while preserving renal function while allowing for a reduction in diuretic dose in some patients.

### In-hospital therapy

Attempts to enhance fluid loss during decongestive therapy generally entails escalating loop diuretic dose or the addition of another diuretic agent.^15^ In the ADVOR trial, adding acetazolamide to loop diuretics resulted in an absolute difference in cumulative diuresis on day 2 of 0.5 liters (95% CI, 0.2–0.8).^23^ In the CLOROTIC trial, adding hydrochlorothiazide to loop diuretics led to weight loss of 1.14 Kg (95% CI, 0.42 to 1.84) at 72 hours.^24^ In this context, daily fluid removal of 678 g in response to AquaPass treatment (operating for ∼4 hours/day) is therefore clinically meaningful; larger amounts can be removed with longer therapy and may be beneficial in some patients.^13^ Thus, the device can provide an alternative to escalation of diuretic therapy or aid in patients with poor response to diuretics or overt diuretic resistance.

### Home therapy

Rates of death and readmission after hospitalization for heart failure remain high despite considerable advances in evidence-based medical treatment.^25–27^ Patients are at highest risk for decompensation requiring readmission in the days and weeks post-hospital discharge,^16^ emphasizing the importance of achieving optimal HF care beyond hospital discharge to decrease avoidable readmissions and improve quality of life. Weight gain occurs in a substantial percentage of patients after hospital discharge. For example, in the ASCEND-HF Trial, 67% of patients showed significant (i.e., more than 5 kg) or moderate (i.e., 1 to 5 kg) weight loss at discharge or day 10; however, at 30 days, only 26% of patients remained with significant or moderate weight loss, whereas 34% showed no weight loss, and 40% experienced weight gain ^27^. An increase of body weight, reflecting a gradual re-accumulation of excess fluid in the post-discharge period, is a major predictor of repeat hospitalization.^27, 28^ While early follow up with volume optimization is recommended by current guidelines,^4, 16^ there are significant knowledge gaps regarding the best downstream post-discharge management strategies.

The dominant role of re-congestion in HF readmissions suggests that current approaches for maintaining euvolemia after hospital discharge can be improved.^29^ While one strategy relies on effective home monitoring to identify physiologic surrogates of fluid retention and early decompensation to avert hospitalization,^30^ corrective actions in ambulatory patients with worsening symptoms of heart failure are limited, centered mainly on increasing the cumulative loop diuretic dose or the addition of a second diuretic. Additionally, current device therapies are designed exclusively for in-hospital use.^31^

In the present study we obtained >2 Kg fluid removal per week with the device on a background of standing diuretic regimen. This resulted in a progressive reduction in congestion score and NT-ProBNP levels with improvements in quality of life. Although the amount of fluid accumulation needed to induce clinical decompensation is variable, it is likely that fluid removal rates achieved in the present study can mitigate or prevent fluid accumulation in the majority of patients, albeit some patients may need more frequent therapies. Our results are also relevant to the goal of safe and effective and more intensive home therapy in outpatients with worsening heart failure as an alternative to hospitalization.^32^

### Advantages of the device

Several device-based strategies have been developed to address congestion in the setting of acute decompensated heart failure, all involving invasive procedures and are therefore targeting the in-hospital phase and cannot be used to maintain decongestion after hospital discharge.^31^ The overall effect of a therapeutic intervention to mitigate congestion also depends on the mechanism of action for fluid removal, the source of the fluid (e.g., intravascular or interstitial) and potential undesirable effects such as neurohormonal activation and worsening renal function. In this context, the AquaPass device provides 2 unique advantages: 1) fluid is removed directly from the interstitial compartment, where most of the excess fluid resides,^33^ thus avoiding rapid intravascular volume changes that may lead to systemic hypotension, neurohormonal activation, reduced renal blood flow and worsening renal function; and 2) the mechanism of action is independent of renal function and diuretic resistance.

### Study Limitations

The results of this study need to be put into the context of several limitations. The number of patients studied was small. There was no standard-of-care comparator arm of diuretic therapy. However, the approach for fluid removal described herein is independent of diuretic efficiency and not intended to enhance diuretic action.

### Conclusion

The current study demonstrates the safety and feasibility of using the AquaPass device to remove fluid in a renal-independent manner in hospitalized ADHF patients, and to promote further weight loss and decongestion following discharge in the home setting. These benefits were achieved without worsening renal function.

## Data Availability

All data produced in the present study are available upon reasonable request to the authors

## Notes

### Competing Interest Statement

Doron Aronson, William T. Abraham, and Daniel Burkhoff are consultants for AquaPass
Yaacov Nitzan is the CEO of AquaPass

### Clinical Trial

NCT05843201

### Funding Statement

The study was funded by AquaPass LTD.

