## Supplemental Data for "Enhancing Sweat Rate for In-Hospital and Home-Based Decongestive Therapy"

**Supplemental Figure 1:** Changes in hourly sweat rate over time in the 12 patients undergoing in-hospital and home therapy. Each graph represents a single patient.

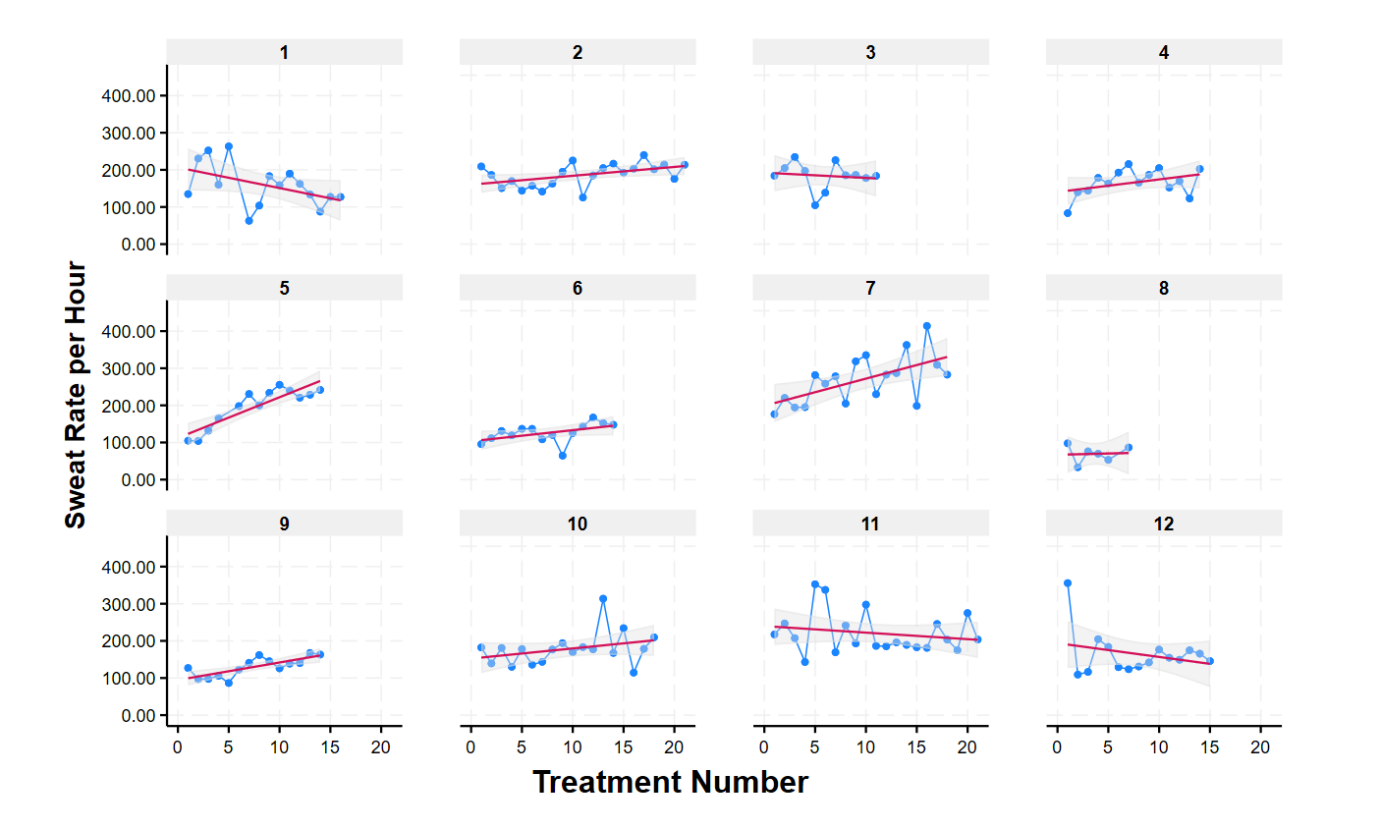

**Supplemental Figure 2:** Weakly Diuretic dose during the hospital phase, hospital discharge and end of home therapy. The line within the box denotes the median and the box spans the interquartile range (25–75th percentiles). Whiskers extend from the 5th to 95th percentiles.

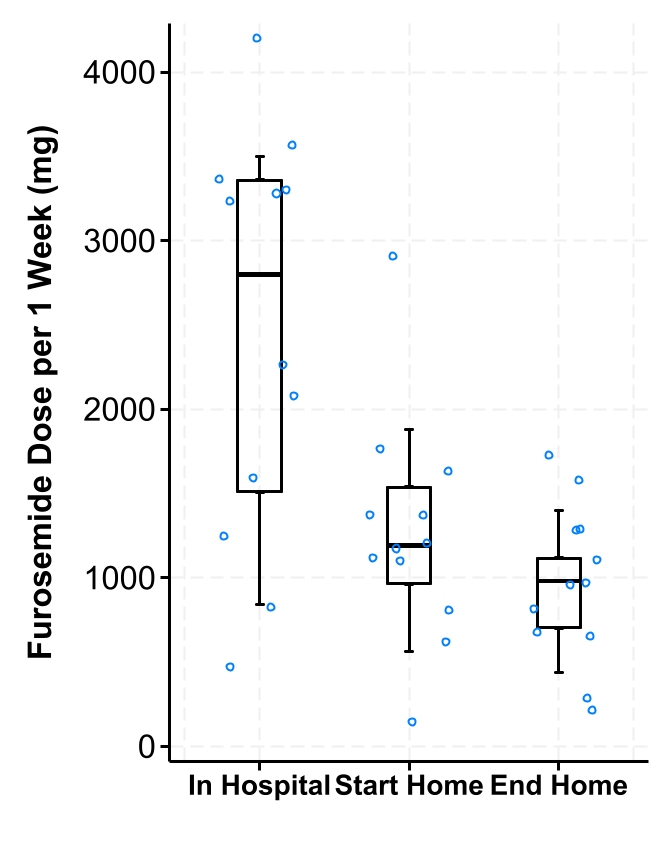

**Supplemental Figure 2:** Relationship between sweat-induced weight loss and estimated glomerular filtration rate (Spearman’s rho 0.04; *P*=0.75).

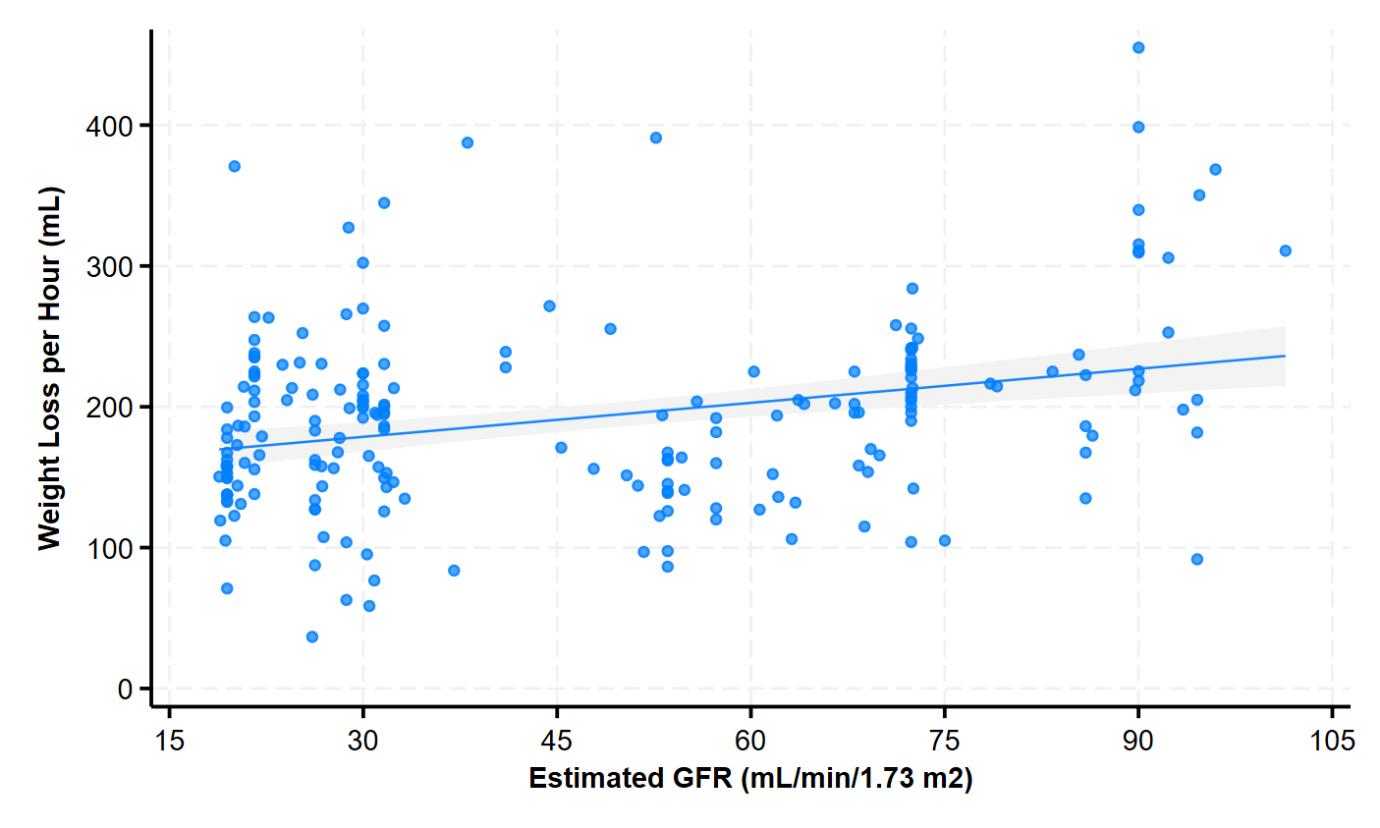
